# External UK validation of the ENDPAC model to predict pancreatic cancer risk: A registered report protocol

**DOI:** 10.1101/2024.05.21.24307690

**Authors:** Claire A. Price, Hugh Claridge, Simon de Lusignan, Natalia Khalaf, Freda Mold, Nadia A. S. Smith, Martyn Winn, Agnieszka Lemanska

**Affiliations:** University of Surrey, School of Health Sciences, Kate Granger Building, 30 Priestley Road, Surrey Research Park, Guildford, Surrey, GU2 7YH, UK; National Physical Laboratory, Hampton Road, Teddington, Middlesex, TW11 0LW, UK; Nuffield Department of Primary Care Health Sciences, University of Oxford, Oxford, OX2 6GG, UK; Center for Innovations in Quality, Effectiveness, and Safety (IQuESt), Michael E. DeBakey Veterans Affairs Medical Center, Baylor College of Medicine Department of Medicine, Houston, TX, USA; Royal Surrey NHS Foundation Trust, Scientific Computing, Egerton Road, Guildford, GU2 7XX, UK; TÜV SÜD UK, Chadwick House, Warrington Road, Birchwood Park, Warrington, WA3 6AE, UK; Scientific Computing Department, Science and Technology Facilities Council, Research Complex at Harwell, Didcot OX11 0FA, UK

**Keywords:** Pancreatic cancer, Routine healthcare data, Early diagnosis, Primary health care, Risk algorithm

## Abstract

**Introduction:** Overall cancer survival has increased over recent decades, but the very low survival rates of pancreatic cancer have hardly changed in the last 50 years. This is attributed to late diagnosis. Pancreatic cancer symptoms are non-specific which makes early diagnosis challenging. Data-driven approaches, including algorithms using combinations of symptoms to predict cancer risk, can aid clinicians. A simple but effective algorithm called Enriching New-Onset Diabetes for Pancreatic Cancer (ENDPAC) has been developed in the United States (US). ENDPAC has not yet been used in the United Kingdom (UK), our aim is to translate ENDPAC into the UK setting. The objectives are to validate ENDPAC and report its predictive utility within primary care.

**Methods:** A retrospective cohort study of people with new-onset diabetes using the nationally representative Oxford-Royal College of General Practitioners Clinical Informatics Digital Hub (ORCHID) database. ORCHID holds over 10 million primary care electronic healthcare records. ENDPAC scores will be calculated for eligible people along with positive predictive value, negative predictive value, sensitivity and specificity of the algorithm. We will evaluate the optimal cut-off for defining people with high-risk of having pancreatic cancer.

**Discussion:** Once validated within the UK, ENDPAC could be implemented in practice to improve early pancreatic cancer diagnosis by using routine data. ENDPAC is currently being tested in the US in a clinical trial to evaluate its effectiveness. ENDPAC offers an automatable and inexpensive way to improve early diagnosis as part of a sequential approach to identify individuals at high-risk of having undiagnosed pancreatic cancer.

**How this fits in:** Pancreatic cancer is a devasting disease which is hard to diagnose. An algorithm called ENDPAC has been developed in the United States to help clinicians identify people at risk of having undiagnosed pancreatic cancer. These people can be referred for an imaging investigation to diagnose or rule out cancer. This protocol outlines a United Kingdom (UK) validation of ENDPAC so that it could be used in clinical practice in the UK.

## Introduction

Pancreatic cancer is a devasting disease due to its high mortality rate with a very low 5-year survival rate of 3-15% [1–6]. It is the 14^th^ most common cancer globally, accounting for 3% of all new cancer cases, with nearly half a million cases worldwide [7]. The high mortality of pancreatic cancer means that it is the fourth major cause of cancer mortality in the world [8].

The need for prompt diagnosis and early detection of pancreatic cancer is widely recognised and is the most likely way in which the current dismal survival rates will improve [9]. However, there are currently no screening or biomarker tests for pancreatic cancer and diagnosis depends upon presentation with symptoms to a General Practitioner (GP). This is challenging as a full-time GP will only see one case every five years on average [10] especially as pancreatic cancer often presents with non-specific symptoms. Symptoms also vary between patients, making diagnosis challenging for clinicians [10–12]. Therefore, it is vital to provide GPs with innovative and practical ways to facilitate early detection. This includes data-driven approaches to flag people who are at high risk of having pancreatic cancer for referrals for further investigation.

A simple but promising data-driven algorithm for the earlier detection of pancreatic cancer has been developed in the United States (US). The Enriching New-onset Diabetes for Pancreatic Cancer (ENDPAC) algorithm [13] uses changes in weight and blood glucose to calculate someone’s risk of having pancreatic cancer. ENDPAC is applied to people aged 50 years or older with new-onset diabetes, as this is the cohort with the highest risk of developing non-familial or sporadic pancreatic cancer [14,15]. This clear clinical rationale means ENDPAC is easily explainable to both GPs and patients. Its relative simplicity and use of measures routinely collected in primary care make ENDPAC well suited for implementation in the UK primary care setting.

One significant advantage of ENDPAC is that it targets early detection as it is based on glucose and weight changes. In contrast, other data-driven risk assessment algorithms [11,16–23] incorporate symptoms like back pain and jaundice, which typically manifest in the later stages of pancreatic cancer. Given that pancreatic cancer is often diagnosed too late for effective treatment, leading to high mortality rates, the ability of ENDPAC to identify cases earlier in the disease progression could potentially save lives. Furthermore, ENDPAC stands out for its use of a relatively small number of routinely collected measurements, addressing concerns about data availability that arise with other algorithms.

Data-driven algorithms, such as Risk Assessment Tools (RATs) like QCancer [24–26] are showing promise in aiding clinical decisions and expediting referrals. However, before ENDPAC can be added to these tools in clinical practice, it requires validation in the UK. ENDPAC was developed in the US and validation in the UK is necessary due to significant differences between the healthcare systems of the two countries [27–30]. Furthermore, previous validations of ENDPAC [21,31,32] have also included secondary care data. Additionally, adjustments to the ENDPAC algorithm are necessary for its application in UK primary care due to how blood glucose is recorded (ENDPAC was developed using fasting blood glucose but in UK primary care, HbA1c (haemoglobin A1c) is recorded and used to diagnose diabetes). Therefore, validation specific to primary care settings in the UK is essential to ensure the algorithm’s suitability for such data.

The aim is to validate the ENDPAC algorithm using UK primary care data. We will extract the cohort of people with new-onset diabetes and their information on pancreatic cancer diagnosis. We will calculate the ENDPAC score at the time of diabetes onset (defined by one HbA1c measure ≥ 48 mmol/mol) reporting predictive value, sensitivity, and specificity of ENDPAC.

## Methods

### Study design and data source

This a retrospective cohort study of adults with new-onset diabetes between 2006 and 2017. They were followed up for 3 years after their first elevated HbA1c reading (defined as diabetes diagnosis) to determine their pancreatic cancer diagnosis status. We used the 2006 cut-off because coding of the metabolic markers such as weight and HbA1c in primary care improved significantly from 2004 with the introduction of the Quality and Outcomes Framework and the pay-for-performance incentive scheme [33,34]. Therefore the 2006 cut-off was set to improve data completeness for at least 2 years before the index date (defined below).

Data from the Oxford-Royal College of General Practitioners Clinical Informatics Digital Hub (ORCHID) database will be used [35]. This is a nationally representative database [36] which downloads electronic healthcare records from GP practices which belong to the Royal College of General Practitioners, Research and Surveillance Centre. In May 2021, using the systematized nomenclature of medicine clinical terminology (SNOMED CT) system [37], we extracted a dataset that included demographic information, ages at diagnoses of diabetes and pancreatic cancer, pancreatic cancer-related symptoms (change in bowel habit, constipation, diarrhoea, abdominal pain, back pain, nausea, vomiting, abdomen scan, operation on pancreas, jaundice, weight loss, indigestion, digestive disorders, suspected pancreatic cancer or cancer referral) as well as all measurements of weight and HbA1c recorded in 2001 or after.

#### Diabetes Status

ENDPAC requires two paired elevated glycaemic values, within 90 days, to diagnose diabetes. However, this requirement is challenging in real-world settings in which people will not necessarily have a second test withing 90 days. Therefore, in this study, diabetes diagnosis is defined by one HbA1c measure of ≥48mmol/mol and encompasses new-onset biochemically-defined hyperglycaemia and diabetes (glycaemic onset). This is the index date.

In the UK HbA1c is measured in mmol/mol (IFCC) [38]. Therefore, HbA1c will be extracted in mmol/mol.

### ENDPAC

The ENDPAC model [13] was developed for people who are over 50 and have developed new-onset diabetes. It combines their age at diabetes diagnosis with changes in their blood glucose and weight over the year before their diabetes diagnosis to produce a score for their risk of having pancreatic cancer. This score consists of three sub-scores: A (Δ blood glucose category) + B (Δ weight category) + C (age category at diabetes diagnosis), see Table 1. The final ENDPAC score produces one of three outcomes: low (<0), intermediate or high (≥3) risk of having pancreatic cancer.

**Table 1:** ENDPAC score parameters with blood glucose (HbA1c) categories equivalent to fasting blood glucose categories presented, based on American Diabetes Association blood glucose levels for healthy, prediabetes, diabetes and uncontrolled diabetes. NOD: New-Onset Diabetes, Δ represents the change in the measurement. Table adapted from Sharma et al. [13]

| Fasting Blood Glucose (FBG) categories | HbA1c Blood Glucose (HbA1c BG) categories | | $\Delta$ BG Category score (NOD – 1y): A |
| --- | --- | --- | --- |
| BG range (mg/dl) | BG range (HbA1c mmol/mol) | Score |  |
| BG category at ~ 1 years |  |  | +1 to +4 |
| < 100 | < 42 | 1 |  |
| $\geq 100$ to < 110 | $\geq 42$ to < 44 | 2 | |
| $\geq 110$ to < 126 | $\geq 44$ to < 48 | 3 | |
| BG category at glycaemically-defined new-onset diabetes |  |  |  |
| $\geq 126$ to < 160 | $\geq 48$ to < 60 | 4 | |
| $\geq 160$ | $\geq 60$ | 5 | |
| $\Delta$ Weight category | | | $\Delta$ Weight score: B |

Table 1: ENDPAC score parameters with blood glucose (HbA1c) categories equivalent to fasting blood glucose categories presented, based on American Diabetes Association blood glucose levels for healthy, prediabetes, diabetes and uncontrolled diabetes.
| $\Delta$ Weight (kg) | Score | Score range |
| --- | --- | --- |
| $\leq -6.0$ | +6 | -6 to +6 |
| $> -6.0$ to $\leq -4.0$ | +4 | |
| $> -4.0$ to $\leq -2.0$ | +2 | |
| $> -2.0$ to $< +2.0$ | 0 | |
| $\geq +2.0$ to $< +4.0$ | -2 | |
| $\geq +4.0$ to $< +6.0$ | -4 | |
| $\geq +6.0$ | -6 | |
| **Age at glycaemically-defined new-onset diabetes** |  | **Age score: C** |
| Age Range | Score | Score range |
| $< 60$ | -1 | -1 to +1 |
| $\geq 60$ to $< 70$ | 0 | |
| $\geq 70$ | +1 | |
| **Total Score** |  | **A + B + C** |

### Patient Population

Study participants will be aged 50 years or older. They will have a diagnosis of new-onset diabetes between 2006 and 2017. The date of diagnosis was defined as the date of the first elevated blood glucose reading (HbA1c ≥ 48 mmol/mol) and defined as the index date. Eligible patients were required to have:

1. An HbA1c ≥48 mmol/mol reading preceded by at least one HbA1c <48 mmol/mol reading in the past 3-24 months
2. No HbA1c ≥48 mmol/mol reading before the index date
3. No history of pancreatic cancer before the index date
4. Weight on the index date (can be ± 30 days)
5. Weight 1 year prior to the index date (between 24 months prior and 6 months prior)

This definition of new-onset diabetes was adapted from the study by Sharma et al. [13]. If multiple readings for weight or HbA1c are available the flowchart by Sharma et al. [36] will be followed to select which readings are used. Sensitivity analysis will then be conducted to investigate the impact of these decisions and whether the predictions change when using highest, lowest or mean readings (when more than one reading is available). Sensitivity analysis will also be conducted on the effect of requiring people to have a second HbA1c reading within 90 days of the first reading as a definition of new-onset diabetes.

### Statistical analysis

#### Descriptive Statistics

Baseline demographic statistics (gender, ethnicity, region and index of multiple deprivation (IMD) quintile) will be extracted.

#### Predictors

The study will include all three risk factors that are part of the ENDPAC model. These are: age at index date, change in body weight (kg), and change in blood glucose (HbA1c).

#### Outcome identification

The outcome is the diagnosis of pancreatic cancer within three years after the index date which is the period of greatest risk of developing pancreatic cancer [14,15,39–41]. Pancreatic cancer diagnosis is based on SNOMED CT system codes, using code 363418001 Malignant tumor of pancreas (disorder) and all sub-codes (excluding 94164003, 94212002, 94325008, 94354002, 94459006, 9460001, 944618007, 285614004) [37]. Excluded codes relate to cancers that have metastasized to the pancreas.

#### Missing data

This study is a validation of the ENDPAC model so ENDPAC scores will only be calculated for people who have the required information (complete cases). Therefore, there will be no missing data.

#### Model validation

The positive predictive value, negative predictive value, sensitivity, specificity as well as the percentage of the cohort to be tested further will be calculated for different cut-off values within the model.

Calibration (predicted probability is in line with observed outcomes) will be assessed using calibration plots of the slope of the observed proportion of events against the predicted risk. The model’s discrimination (ability to distinguish between those with and without the pancreatic cancer outcome) will be assessed by area under the receiver operating characteristic curve.

The optimal cut-off value to identify people at high-risk of pancreatic cancer will then be found, using the Youden index, which maximises both the sensitivity and specificity of the model [42].

### Subgroup analyses

Risk score stratification will be utilised to explore the differences in cancer rates between the risk score categories (low-, intermediate- and high-risk). The proportion of people who would be referred for further testing will also be calculated (overall and when data is stratified). Model sensitivity will also be investigated for different lead times (3-6 months, 6-9 months, 9-15 months, 15-18 months, 18-21 months and 21-24 months) of the measurements utilised. The effect of using a single elevated HbA1c reading as the definition of diabetes instead of paired values will also be investigated.

### Software and reproducibility

The database is managed in Structured Query Language (SQL) Server Management Studio version v18.9.1. Data analyses will be conducted using RStudio version 2023.03.0+386 “Cherry Blossom”. Software will be published open access via GitHub for reuse and review.

## Data Availability

Data will remain under the control of the Oxford-Royal College of General Practitioners Clinical Informatics Digital Hub (ORCHID, orchid.phc.ox.ac.uk) and can be accessed following all necessary approvals.

## Ethical Implications

The study was approved by the University of Surrey Ethics Committee (reference number: FHMS 21-22 269 EGA). Access to ORCHID data has been approved by Royal College of General Practitioners (RCGP) Research and Surveillance Centre (RSC) under data request RSC_0420.

## Dissemination

Dissemination is planned through peer-reviewed publications and conference presentations.

## Acknowledgements

We thank patients and practices who are members of the ORCHID network. This project was funded as part of an EPSRC iCase studentship undertaken by CP. The work of NPL co-authors was funded by the UK Government’s Department for Science, Innovation & Technology through the UK’s National Measurement System programmes.

## Notes

### Competing Interest Statement

The authors have declared no competing interest.

### Author Declarations

The Ethics Committee of University of Surrey gave ethical approval for this work (reference number: FHMS 21-22 269 EGA). Access to ORCHID data has been approved by Royal College of General Practitioners (RCGP) Research and Surveillance Centre (RSC) under data request RSC_0420.

## References

[1] Rachet B, Maringe C, Nur U, Quaresma M, Shah A, Woods LM, et al. Population-based cancer survival trends in England and Wales up to 2007: an assessment of the NHS cancer plan for England. Lancet Oncol 2009;10:351–69.

[2] Rahib L, Smith BD, Aizenberg R, Rosenzweig AB, Fleshman JM, Matrisian LM. Projecting cancer incidence and deaths to 2030: the unexpected burden of thyroid, liver, and pancreas cancers in the United States. Cancer Res 2014;74:2913–21.

[3] Woodmansey C, McGovern AP, McCullough KA, Whyte MB, Munro NM, Correa AC, et al. Incidence, Demographics, and Clinical Characteristics of Diabetes of the Exocrine Pancreas (Type 3c): A Retrospective Cohort Study. Diabetes Care 2017;40:1486–93.

[4] Bray F, Ferlay J, Soerjomataram I, Siegel RL, Torre LA, Jemal A. Global cancer statistics 2018: GLOBOCAN estimates of incidence and mortality worldwide for 36 cancers in 185 countries. CA Cancer J Clin 2018;68:394–424.

[5] Arnold M, Rutherford MJ, Bardot A, Ferlay J, Andersson TM-L, Myklebust TÅ, et al. Progress in cancer survival, mortality, and incidence in seven high-income countries 1995-2014 (ICBP SURVMARK-2): a population-based study. Lancet Oncol 2019;20:1493–505.

[6] Siegel RL, Miller KD, Wagle NS, Jemal A. Cancer statistics, 2023. CA Cancer J Clin 2023;73:17–48.

[7] Sung H, Ferlay J, Siegel RL, Laversanne M, Soerjomataram I, Jemal A, et al. Global Cancer Statistics 2020: GLOBOCAN Estimates of Incidence and Mortality Worldwide for 36 Cancers in 185 Countries. CA Cancer J Clin 2021;71:209–49.

[8] Wang W, Chen S, Brune KA, Hruban RH, Parmigiani G, Klein AP. PancPRO: risk assessment for individuals with a family history of pancreatic cancer. J Clin Oncol 2007;25:1417–22.

[9] Kenner BJ, Chari ST, Maitra A, Srivastava S, Cleeter DF, Go VLW, et al. Early Detection of Pancreatic Cancer-a Defined Future Using Lessons From Other Cancers: A White Paper. Pancreas 2016;45:1073–9.

[10] Evans J, Chapple A, Salisbury H, Corrie P, Ziebland S. “It can’t be very important because it comes and goes”—patients’ accounts of intermittent symptoms preceding a pancreatic cancer diagnosis: a qualitative study. BMJ Open 2014;4:e004215.

[11] Hippisley-Cox J, Coupland C. Identifying patients with suspected pancreatic cancer in primary care: derivation and validation of an algorithm. Br J Gen Pract 2012;62:e38–45.

[12] Olson SH, Xu Y, Herzog K, Saldia A, DeFilippis EM, Li P, et al. Weight Loss, Diabetes, Fatigue, and Depression Preceding Pancreatic Cancer. Pancreas 2016;45:986–91.

[13] Sharma A, Kandlakunta H, Nagpal SJS, Feng Z, Hoos W, Petersen GM, et al. Model to Determine Risk of Pancreatic Cancer in Patients With New-Onset Diabetes. Gastroenterology 2018;155:730-739.e3.

[14] Chari ST, Leibson CL, Rabe KG, Ransom J, de Andrade M, Petersen GM. Probability of pancreatic cancer following diabetes: a population-based study. Gastroenterology 2005;129:504–11.

[15] Pannala R, Basu A, Petersen GM, Chari ST. New-onset diabetes: a potential clue to the early diagnosis of pancreatic cancer. Lancet Oncol 2009;10:88–95.

[16] Collins GS, Altman DG. External validation of QDSCORE(®) for predicting the 10-year risk of developing Type 2 diabetes. Diabet Med 2011;28:599–607.

[17] Boursi B, Finkelman B, Giantonio BJ, Haynes K, Rustgi AK, Rhim AD, et al. A clinical prediction model to assess risk for pancreatic cancer among patients with prediabetes. Eur J Gastroenterol Hepatol 2022;34:33–8.

[18] Boursi B, Finkelman B, Giantonio BJ, Haynes K, Rustgi AK, Rhim AD, et al. A Clinical Prediction Model to Assess Risk for Pancreatic Cancer Among Patients With New-Onset Diabetes. Gastroenterology 2017;152:840-850.e3.

[19] Baecker A, Kim S, Risch HA, Nuckols TK, Wu BU, Hendifar AE, et al. Do changes in health reveal the possibility of undiagnosed pancreatic cancer? Development of a risk-prediction model based on healthcare claims data. PLoS One 2019;14:e0218580.

[20] Appelbaum L, Cambronero JP, Stevens JP, Horng S, Pollick K, Silva G, et al. Development and validation of a pancreatic cancer risk model for the general population using electronic health records: An observational study. Eur J Cancer 2021;143:19–30.

[21] Chen W, Butler RK, Lustigova E, Chari ST, Wu BU. Validation of the Enriching New-Onset Diabetes for Pancreatic Cancer Model in a Diverse and Integrated Healthcare Setting. Dig Dis Sci 2021;66:78–87.

[22] Chen W, Zhou B, Jeon CY, Xie F, Lin Y-C, Butler RK, et al. Machine learning versus regression for prediction of sporadic pancreatic cancer. Pancreatology 2023;23:396–402.

[23] Malhotra A, Rachet B, Bonaventure A, Pereira SP, Woods LM. Can we screen for pancreatic cancer? Identifying a sub-population of patients at high risk of subsequent diagnosis using machine learning techniques applied to primary care data. PLoS One 2021;16:e0251876.

[24] Price S, Spencer A, Medina-Lara A, Hamilton W. Availability and use of cancer decision-support tools: a cross-sectional survey of UK primary care. Br J Gen Pract 2019;69:e437–43.

[25] Akanuwe JNA, Black S, Owen S, Siriwardena AN. Barriers and facilitators to implementing a cancer risk assessment tool (QCancer) in primary care: a qualitative study. Prim Health Care Res Dev 2021;22:e51.

[26] Chiang PP-C, Glance D, Walker J, Walter FM, Emery JD. Implementing a QCancer risk tool into general practice consultations: an exploratory study using simulated consultations with Australian general practitioners. Br J Cancer 2015;112 Suppl 1:S77–83.

[27] Ham C. Money can’t buy you satisfaction. BMJ 2005;330:597–9.

[28] Starfield B. Why is the grass greener? BMJ 2005;330:727–9.

[29] Docteur E, Berenson RA. Timely analysis of immediate health policy issues. Urban Institute 2009. https://www.urban.org/sites/default/files/publication/30596/411947-How-Does-the-Quality-of-U-S-Health-Care-Compare-Internationally-.PDF (accessed 8 April 2023).

[30] Thompson L. A Comparative Evaluation of the Structure of Primary Care In the United States and the United Kingdom. 2010.

[31] Khan S, Safarudin RF, Kupec JT. Validation of the ENDPAC model: Identifying new-onset diabetics at risk of pancreatic cancer. Pancreatology 2021;21:550–5.

[32] Boursi B, Patalon T, Webb M, Margalit O, Beller T, Yang Y-X, et al. Validation of the enriching new-onset diabetes for pancreatic cancer model. Pancreas 2022;51:196–9.

[33] NHS Digital. National Quality and Outcomes Framework. NHS Digital 2005. https://digital.nhs.uk/data-and-information/publications/statistical/quality-and-outcomes-framework-achievement-prevalence-and-exceptions-data/national-quality-and-outcomes-framework-statistics-for-england-2004-05 (accessed 3 March 2023).

[34] McGovern A, Hinton W, Correa A, Munro N, Whyte M, de Lusignan S. Real-world evidence studies into treatment adherence, thresholds for intervention and disparities in treatment in people with type 2 diabetes in the UK. BMJ Open 2016;6:e012801.

[35] de Lusignan S, Liyanage H, Hobbs R. Health Data Research Innovation Gateway. ORCHID Clinical Informatics Digital Hub 2021. https://web.www.healthdatagateway.org/collection/5626663352808625 (accessed 22 March 2023).

[36] Correa A, Hinton W, McGovern A, van Vlymen J, Yonova I, Jones S, et al. Royal College of General Practitioners Research and Surveillance Centre (RCGP RSC) sentinel network: a cohort profile. BMJ Open 2016;6:e011092.

[37] NHS Digital. Snomed-CT 2017. https://termbrowser.nhs.uk/?perspective=full&conceptId1=363418001&edition=uk-edition&release=v20231122&server=https://termbrowser.nhs.uk/sct-browser-api/snomed&langRefset=999001261000000100,999000691000001104 (accessed 12 December 2023).

[38] HbA1c reporting methods set to change. Diabetes UK 2011. https://www.diabetes.org.uk/about_us/news_landing_page/hba1c-reporting-methods-set-to-change (accessed 21 September 2023).

[39] Andersen DK, Korc M, Petersen GM, Eibl G, Li D, Rickels MR, et al. Diabetes, Pancreatogenic Diabetes, and Pancreatic Cancer. Diabetes 2017;66:1103–10.

[40] Pannala R, Leirness JB, Bamlet WR, Basu A, Petersen GM, Chari ST. Prevalence and clinical profile of pancreatic cancer-associated diabetes mellitus. Gastroenterology 2008;134:981–7.

[41] Aggarwal G, Rabe KG, Petersen GM, Chari ST. New-onset diabetes in pancreatic cancer: a study in the primary care setting. Pancreatology 2012;12:156–61.

[42] Fluss R, Faraggi D, Reiser B. Estimation of the Youden Index and its associated cutoff point. Biom J 2005;47:458–72.

